# Bayesian modeling of COVID-19 cases with a correction to account for under-reported cases

**DOI:** 10.1101/2020.05.24.20112029

**Authors:** Anderson Castro Soares de Oliveira, Lia Hanna Martins Morita, Eveliny Barroso da Silva, Daniele Cristina Tita Granzotto, Luiz André Ribeiro Zardo, Cor Jesus Fernandes Fontes

## Abstract

The novel of COVID-19 disease started in late 2019 making the worldwide governments came across a high number of critical and death cases, beyond constant fear of the collapse in their health systems. Since the beginning of the pandemic, researchers and authorities are mainly concerned with carrying out quantitative studies (modeling and predictions) overcoming the scarcity of tests that lead us to under-reporting cases. To address these issues, we introduce a Bayesian approach to the SIR model with correction for under-reporting in the analysis of COVID-19 cases in Brazil. The proposed model was enforced to obtain estimates of important quantities such as the reproductive rate and the average infection period, along with the more likely date when the pandemic peak may occur. Several under-reporting scenarios were considered in the simulation study, showing how impacting is the lack of information in the modeling.

## 1. Introduction

The COVID-19 epidemic disease is caused by the new SARS-CoV-2 coronavirus associated with the severe acute respiratory syndrome (SARS) that began in Wuhan, China, late 2019 (Rodríguez-Morales et al., 2020). After the first detected case in China, the disease continued to spread globally with exported cases confirmed in all of the continents worldwide. In a matter of a few months, the disease overtook 80 thousand reported cases until early April, 2020. On March 12nd, 2020 the World Health Organization (WHO) declared COVID-19 as pandemic disease, when more than 20 thousand cases and almost a thousand deaths were registered in the European Region - the center of this pandemic according to the Europe’s Standing Committee (WHO, 2020).

There are still many unknowns about COVID-19 and the lack of evidence complicates the design of appropriate response policies - for example, it is impossible to precisely say something about the mortality rate and determine the disease recurrence rate (Lenzer, 2020).

Despite uncertainties, the frightening speed through which this disease spreads across communities and the collapse that it is capable of causing to the health systems are facts that must be faced. The exponential growth of the cases and the consequent number of deaths had been observed in a short period. In mid-January 2020, a few weeks after the first detected COVID-19 case in the world, the countries that are close to the territory of the virus origin, on the Asian continent, as well in European and American Regions also began to report cases of the disease. Five months later, more than 200 countries and territories around the world have reported over to 3 million confirmed cases of COVID-19 and a death toll of about 200 thousand people.

In Brazil, the first confirmed COVID-19 case occurred on February 25th, 2020. This first case was a 61 years-old male, who stayed from February 9th to February 20th, 2020 in Lombardy - an Italian region were a significant outbreak was ongoing at that time. On March 17th, the health authorities in São Paulo confirmed the Brazilian death from the new coronavirus. The victim, whose identity has not been disclosed, had been hospitalized in São Paulo city.

Preserving due proportions, COVID-19 is not the first experienced significant outbreaks of infections that were declared Public Health Emergencies of International Concern by the WHO. Year after year we also have experimented with the Zika and Chikungunya outbreaks in the last decade and continue facing the huge consequences of dengue. Confronting outbreaks in the large Brazilian territory is a twofold problem. The first is the demographic and territorial size of the country, with an estimated population of 210 million according to the Brazilian Institute for Geography and Statistics and the heterogeneity intrinsic to its extensive territory. Another problem pointed out by the past epidemics run into a recurring problem of under-reporting (de Oliveira et al., 2017; Stoner et al., 2019).

The COVID-19, given its complexity and behavior, exposed the problem of under-reporting disease occurrence not only in Brazil but in several countries worldwide. As a consequence, the lack of information has launched a warning about the researchers of the world concerning models and estimates, since the database available may not be reliable from what had indeed been observed.

Focusing on the modeling and estimating, aiming to preview the behavior and the speed of the COVID-19 growth, this paper presents an approach to address the problem of under-registration of COVID-19 cases in Brazil, proposing methodologies to work on the inaccuracy of the official reported cases. Then, we investigate a general framework for correcting under-reporting data making it possible to perform a model, in a Bayesian framework, which allows great flexibility and leads to complete predictive distributions for the true counts, therefore quantifying the uncertainty in correcting the under-reporting. Several scenarios of under-reporting were considered in a simulation study, presenting the real lack of data impact.

This paper is organized as follows. Section 2 describes the methodology for estimating the reported rates. In Section 3, we introduce the SIR model for modeling epidemics. In Section 4, we introduce the Bayesian framework for the SIR model with a modification to account for under-reporting. In Section 5 we show the model application for COVID-19 cases in Brazil and in Section 6, we present a simulation study of the proposed model. Finally, in Section 7, we give some concluding remarks.

## 2. Reported rate estimation

Although in the first moment there was a real hunt for the size and the moment of the COVID-19 cases peak, the most important aspects of the outbreak are the growth rate of the infection. Statistical and mathematical models are being used to preview the rates and analyze the growth curve behavior to assist health public managers in decision-making (Cotta et al., 2020).

According to Kim et al. (2020), estimating the case fatality rate (CFR) is a high priority in response to this pandemic. This fatality rate is the proportion of deaths among all confirmed patients with the disease, which has been used to assess and compare the severity of the epidemic between countries. The rates can also be used to assess the healthcare capacity in response to the outbreak. Indeed, several researchers are interested in estimating the CFR in the peak of the outbreak, analyzing its variation among different countries, and check the influence of other features as ages, gender, and physical characteristics in the CFR of the COVID-19.

Aiming to estimate the CFR, first of all, lets set up the Brazilian scenario of COVID-19 case notification: the Brazilian Ministry of Health collects daily all confirmed cases data for Brazil and all its states. Although the data presented by the health authorities are official, they are only from patients with COVID-19 confirmed by blood and/or swab positive tests. Given the scarcity of tests for all the suspected individuals, the notified patients are only those with severe disease or that demanding hospitalization. It is relevant to highlight that no clinically diagnosed patient, even those with symptoms compatible with the disease have been officially counted, evidencing an under-reporting of the case frequency.

Faced with the lack of COVID-19 tests, which naturally leads to the underreporting data, before any modeling purpose we have the desire to correct and update the current numbers, bringing them as close as possible to reality.

Following Russel et al. (2020), we also based on a delay-adjusted case fatality ratio to estimate under-reporting, using the incidence of cases and deaths to estimate the number of notified cases by

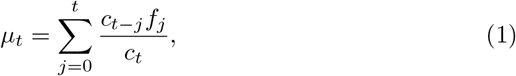

where *c_t_* is the daily incidence of cases at the moment *t, f_j_* is the proportion of cases with a delay between the confirmation and the death, and *μ_t_* represents the underestimation proportion of cases with known outcomes, (Nishiura et al., 2009).

Then, the corrected CFR is given by

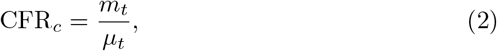

where *m_t_* is the cumulative number of deaths.

To estimate the potential for under-reporting, we assume that the CFR is 1.4% with a 95% confidence interval from 1.2% up to 1.7% found in China (Guan W-j, 2020). Thus, the potential for reporting rate is given by

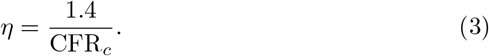

## 3. The SIR Model

Epidemic models are tools widely used to study the mechanisms by which diseases spread, to predict the course of an outbreak, and to evaluate strategies to control an epidemic disease. Several analyses of an epidemic spreading disease can be found in the literature that applies the time series model (given the historical data), the log-logistic family of models (the Chapman, Richards, among others), and compartments models (Bjørnstad, 2018).

Kermack & McKendrick (1927) proposed a class of compartmental models that simplified the mathematical modeling of infectious disease transmission. Entitled as SIR model, it is a set of general equations which explains the dynamics of an infectious disease spreading through a susceptible population. Essentially, the standard SIR model is a set of differential equations that can suit the Susceptible (if previously unexposed to the pathogen), Infected (if currently colonized by the pathogen), and Removed (either by death or recovery) as follows:

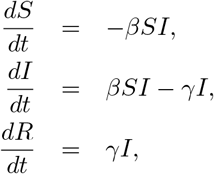

where *S*, *I* and *R* are the total number of susceptible, infected and removed individuals in the population, respectively, *γ* is the removal rate and *β* is the infectious contact rate.

It is important to note that

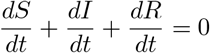

and so, the total population, *S*(*t*) + *I*(*t*) + *R*(*t*) remains constant for all *t* ≥ 0.

For the practical point of view, the most interesting issue is to estimate 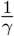, which determines the average infection period and the basic reproductive ratio 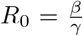, defined as the expected number of secondary infections from a single index case in a completely susceptible population (Keeling & Rohani, 2011).

## 4. Bayesian Approach

The Bayesian methods are used in several works (Gelman et al., 1995); (Paulino et al., 2018). The Bayesian approach in the context of the SIR model is a flexible way to account for uncertainty in the parameters, in the form of the disease transmission dynamic. The Dirichlet-Beta state-space model appears in some papers as Osthus et al. (2017) and Song et al. (2020). The target distribution for inference is the *a posteriori* distribution of the quantities of interest, more specifically *β*, *γ*, and *R*_0_: the infectious contact rate, the removal rate, and the propagation rate, respectively. The application of this methodology is through Markov chain Monte Carlo methods (MCMC) through Gibbs Sampling and the Metropolis-Hastings algorithm (Chib & Greenberg, 1995).

The use of Dirichlet distribution for the proportions of susceptible, infected, and removed individuals in the target population are a feasible way to guarantee that the support set of these quantities has boundaries, for example, the number of infected individuals must be always positive.

### 4.1. Model specification

In this section, we present a modification to account for under-reporting in the context of the Dirichlet-Beta state-space model from Osthus et al. (2017). This adaptation is based on a reparametrization of Beta distribution that includes the reported rate estimate, *η*, from equation (3).

The Beta distribution, as is well known, is very flexible for proportions modeling since its density can have quite different shapes depending on the values of the two parameters that index this distribution (Ferrari & Cribari-Neto, 2004). For this reason, we made a reparametrization to the Beta model in such a way that we could obtain a regression structure for the means of the response variables associated with a precision parameter.

Let 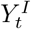 be the reported infected proportion, 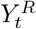 be the reported removed proportion and 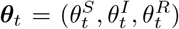 be the true but unobservable susceptible, infectious, and removed proportions of the population, respectively.

Hence, we rewrite the SIR model in terms of these unobservable proportions as the following

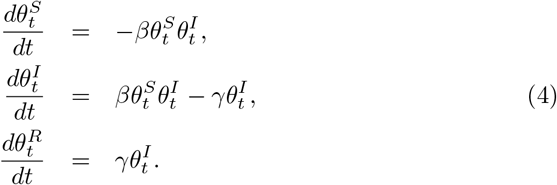

Then, the distributions for 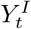, 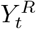, and ***θ****_t_* are given below

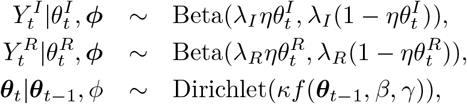

where *ϕ* = (*β*, *γ*, ***θ***_0_, *κ*, *λ*) is the parameter vector for this model and *f* (***θ****_t_*_−1_, *β*, *γ*) is the solution for the differential equations in (4).

Note that it is necessary to obtain the solutions for the proportions 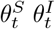 and 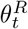. These solutions can be found using the Runge-Kutta fourth-order method, in short RK4, for solving non-linear ordinary differential equations (Mathews, 1992) and can be seen in Appendix A.

## 5. Case study: the COVID-19 Brazilian data

The official Brazilian data consists of daily collections carried out by the national health department with records of infected individuals and deaths in all states and national territory, from February 26th, 2020 when the first case of COVID-19 was registered up to May 20th, 2020.

It is notable in Brazil a lack of testing due to the registry of only severe cases and consequently under-reporting cases of COVID-19. Taking this fact into account, we consider for this research not only the official data but also the estimates of reported rate.

### 5.1. Reported Rate of COVID-19

In order to obtain the estimate of reported rate, assume that the delay in confirmation until death follows the same estimated distribution of hospitalization until death. Using data from COVID-19 in Wuhan, China, between December 17th, 2019, and January 22nd, 2020, it has a lognormal distribution with mean of 13, median of 9.1 and standard deviation of 12.7 days (Linton NM, 2020). This methodology based on the information of delay from hospitalization until death is reasonable since China was considered as one of the countries that most tested the population for the virus, and consequently, it is supposed to have a tiny under-reporting rate.

Using the methodology presented in section 2, the reporting rate in Brazil, *η*, was estimated to be 0.07 with 95% confidence interval from 0.06 up to 0.08. Prado et al. (2020) obtained a reporting rate of 0.08 with data from Brazil until April 10th, 2020. These results are similar to the analysis from Ribeiro & Bernardes (2020), which present a 7.7: 1 under-reporting rate, meaning that the real cases in Brazil should be, at least, seven times the published number.

Table 1 presents the rates for all states of Brazil, from which we can observe that Paralba has the lowest reported rate 0.06 and while Roraima presents the highest reported rate 0.52. Indeed, Prado et al. (2020) found that Paraíba and Pernambuco had a low reporting rate comparing with other states.

**Table 1:** Reported rate estimates and 95% confidence interval (95% CI) for COVID-19 Brazilian data.

| State | Rate ($\hat{\eta}$) | Lower 95% CI | Upper 95% CI |
| --- | --- | --- | --- |
| Acre | 0.14 | 0.12 | 0.17 |
| Alagoas | 0.10 | 0.09 | 0.12 |
| Amapa | 0.20 | 0.17 | 0.24 |
| Amazonas | 0.08 | 0.07 | 0.10 |
| Bahia | 0.20 | 0.17 | 0.24 |
| Ceará | 0.11 | 0.10 | 0.13 |
| Distrito Feral | 0.40 | 0.34 | 0.48 |
| Espírito Santo | 0.19 | 0.16 | 0.23 |
| Goiás | 0.19 | 0.17 | 0.24 |
| Maranhão | 0.11 | 0.09 | 0.13 |
| Mato Grosso | 0.22 | 0.19 | 0.27 |
| Mato Grosso do Sul | 0.24 | 0.21 | 0.30 |
| Minas Gerais | 0.19 | 0.16 | 0.23 |
| Pará | 0.10 | 0.08 | 0.12 |
| Paraíba | 0.06 | 0.06 | 0.08 |
| Paraná | 0.15 | 0.13 | 0.18 |
| Pernambuco | 0.07 | 0.06 | 0.09 |
| Piauí | 0.10 | 0.09 | 0.12 |
| Rio de Janeiro | 0.08 | 0.07 | 0.10 |
| Rio Grande do Norte | 0.14 | 0.12 | 0.17 |
| Rio Grande do Sul | 0.23 | 0.20 | 0.28 |
| Rondônia | 0.17 | 0.15 | 0.21 |
| Roraima | 0.52 | 0.44 | 0.63 |
| Santa Catarina | 0.30 | 0.25 | 0.36 |
| São Paulo | 0.09 | 0.07 | 0.10 |
| Sergipe | 0.12 | 0.10 | 0.14 |
| Tocantins | 0.17 | 0.15 | 0.21 |

### 5.2. Estimation: Dirichlet-Beta state-space model

For the adjustment of the Bayesian model, the *prioris* and hyper-parameters are specified:

- *γ* - We assume that the average infection period is equal to 15 days. Thus, the *γ a priori* belongs to lognormal distribution with mean of 0.07 and variance of 0.01.

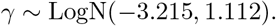 The average infection period *ρ* comes directly from *γ* parameter, that is, 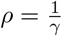.
- *β* - The reproduction number *R*_0_ of the disease is estimated by the ratio 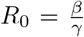 We assume that *R*_0_ *a priori* belongs to lognormal distribution with mean of 3 and variance of 9. Thus *β* values were obtained from *β* = R_0_*_γ_*

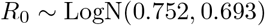
- The *a priori* distributions for *κ*, *λ_I_* and *λ_R_* and ***θ***_0_ were obtained according to Osthus et al. (2017), that is,

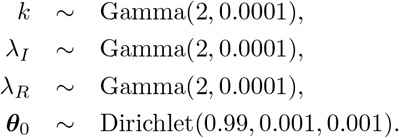

The estimates from *a posteriori* distributions for *R*_0_, *β*, *γ*, *κ*, *λ_I_* and *λ_R_* were obtained through MCMC methods, specifically Gibbs sampling (Geman & Geman, 1984). To execute the sampling procedure, we used the R programming language (R Core Team, 2020), with rjags package (Plummer, 2019).

The total number of iterations considered, as well as the discard (burn-in) and the minimum distance between one iteration to another (thin) were obtained through the criterion of Raftery & Lewis (1992) in the analysis of a pilot sample with 10,000 iterations.

The convergence diagnosis of the MCMC procedure was verified using the Geweke (Geweke, 1992) and Heidelberger and Welch(Heidelberger & Welch, 1983) criteria, which are available in the coda package (Plummer et al., 2006).

Table 2 shows the p-values from Geweke, and Heidelberger and Welch convergence diagnostics, from which we conclude that chains reached convergence for all parameters (p-value > 0.05). The inference was made by considering the reported rate estimate in Brazil, 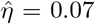, a chain of 300,000 interactions was generated, with a burn-in of 10,000 and a thin of 300, resulting in a final sample of 1,450 values.

**Table 2:** P-values for Geweke, and Heidelberger and Welch convergence diagnostics.

| parameter | Geweke | Heidelberger and Welch |
| --- | --- | --- |
| $R_0$ | 0.7222 | 0.2026 |
| $\beta$ | 0.8900 | 0.1898 |
| $\gamma$ | 0.8210 | 0.2611 |
| $\kappa$ | 0.2965 | 0.1455 |
| $\lambda_I$ | 0.8205 | 0.2462 |
| $\lambda_R$ | 0.1118 | 0.2568 |

The parameter estimates are shown in Table 3, in which 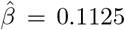 and 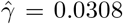 are the major characteristics from SIR model and 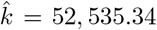, 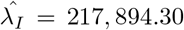 and 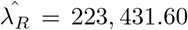 express the magnitude of the process error for the unknown proportions (***θ***) in Bayesian approach.

**Table 3:** Point estimates and 95% Credible Interval.

| parameter | Mode | 95% Credible Interval |  |
| --- | --- | --- | --- |
|  |  | lower | upper |
| $\gamma$ | 0.0308 | 0.0272 | 0.0343 |
| $\beta$ | 0.1125 | 0.1067 | 0.1201 |
| $R_0$ | 3.6243 | 3.3528 | 4.0335 |
| $\rho$ | 32.1667 | 29.1268 | 36.7576 |
| $\kappa$ | 52535.34 | 38384.26 | 71244.52 |
| $\lambda_I$ | 217894.30 | 148822.20 | 310111.60 |
| $\lambda_R$ | 223431.60 | 147997.80 | 320880.00 |

The inference results show that 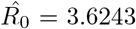 which expresses a high reproductive rate of the virus. Also, 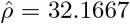 days shows that the time for virus infection is very close to one month period.

Using the parameter estimates from Table 3 and the latent proportion (***θ***), we reached information about the peak from SIR curve for the COVID-19 transmission in Brazil, that is the time when the proportion of infected individuals reaches its maximum. The peak estimate is June 18th, 2020, occurring between June 12nd and June 22th, 2020 and it is shown in Figure 1.

**Figure 1:**
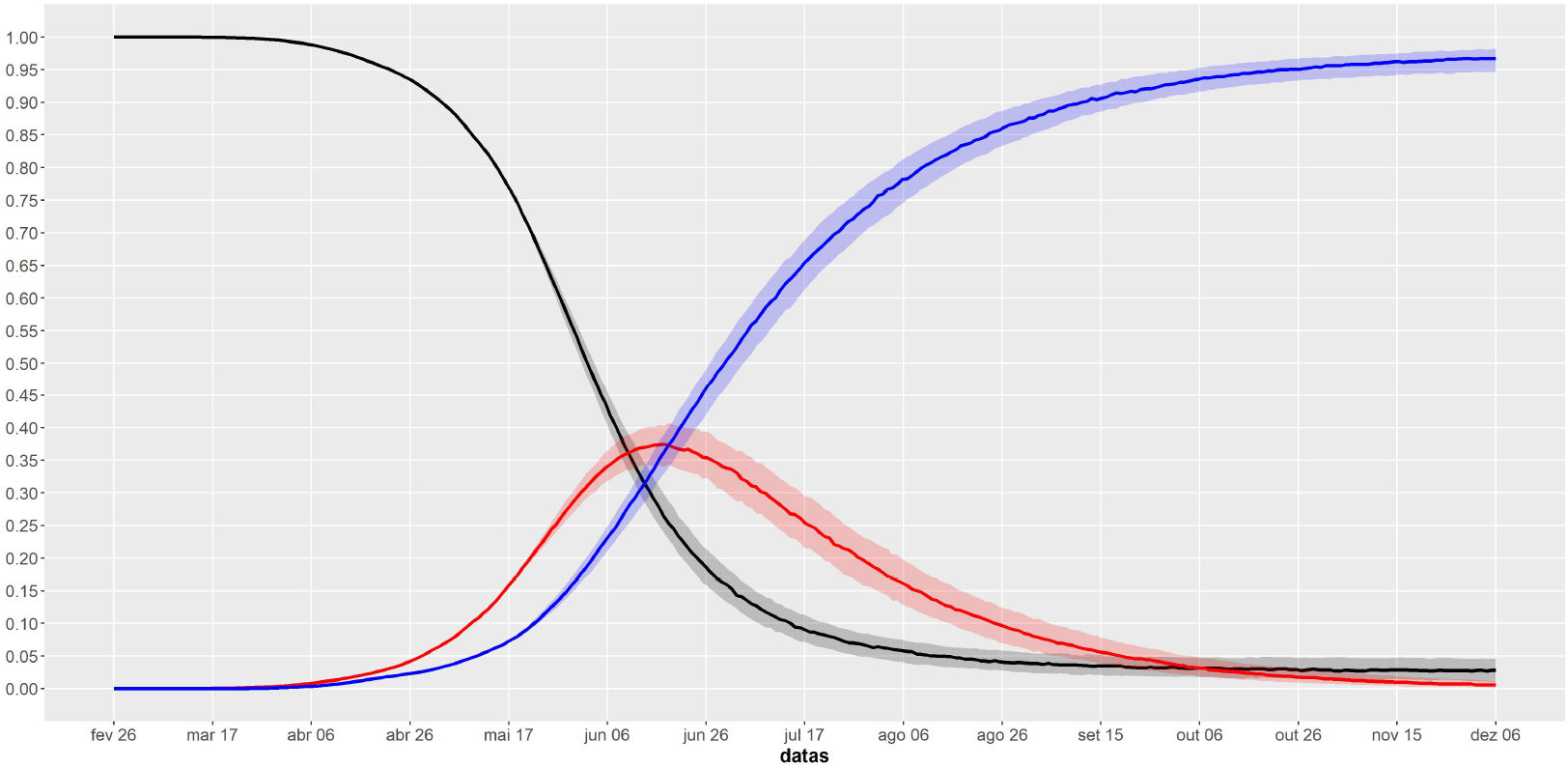
Estimated SIR curves for COVID-19 Brazilian data from February 26th to May 20th, 2020.

## 6. Simulation Study

Concerning to evaluate the effect of the notification rate on the model’s estimates, a simulation study was carried out. The model was estimated considering COVID-19 data in Brazil, assuming a reporting rate between 0.05 and 1.00, varying every 0.05. Aiming the practical point of view, we conduct a simulation study to investigate the effects of under-reporting in the parameters of the SIR model and how it impacts on the pandemic curve behavior. For each value of *η*, a chain of 300,000 interactions was generated, with a burn-in of 10,000 and a thin of 300.

Figure 2 shows the point estimates and 95% credible intervals for *β* and *γ* versus the reported rate values. It can be observed that as reported rate increases, *β* estimate becomes lower, which means that the infectious contact rate is underestimated when under-reporting is ignored. Additionaly, the removal rate *γ* remains almost constant when the reported rate increases, which means that it is not influenced by the rates.

**Figure 2:**
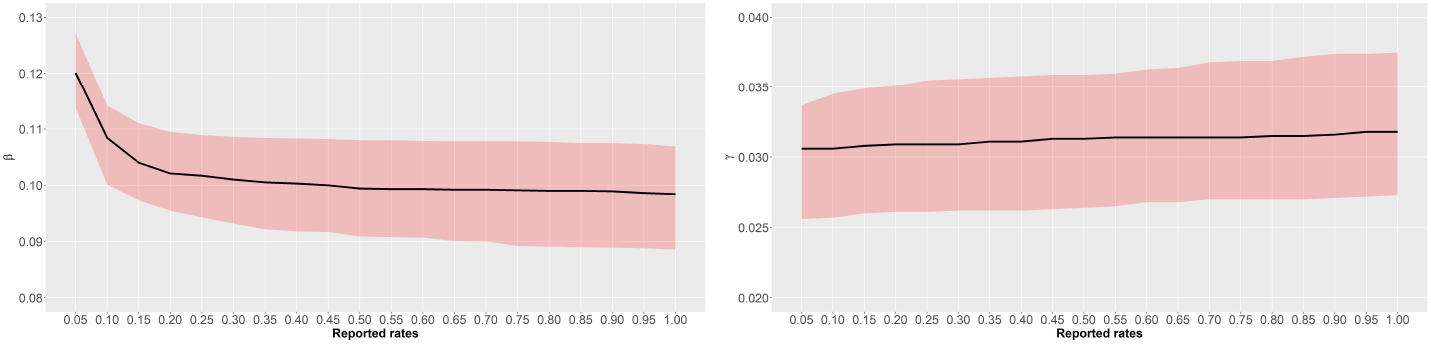
Point estimates and 95% credible intervals for *β* and *γ* versus reported rates

The graphics with the point estimates and 95% credible intervals for *R*_0_ and infection period ρ versus the reported rates are shown in Figure 3, from which we observe that *R*_0_ decreases as the reported rate increases and ρ keeps roughly invariant, then we can conclude that the reproduction rate and infection period can be underestimated when under-reporting is ignored, affording an unreal impression on a tiny mean number of secondary individuals that a primary individual can infect, when in fact it is large.

**Figure 3:**
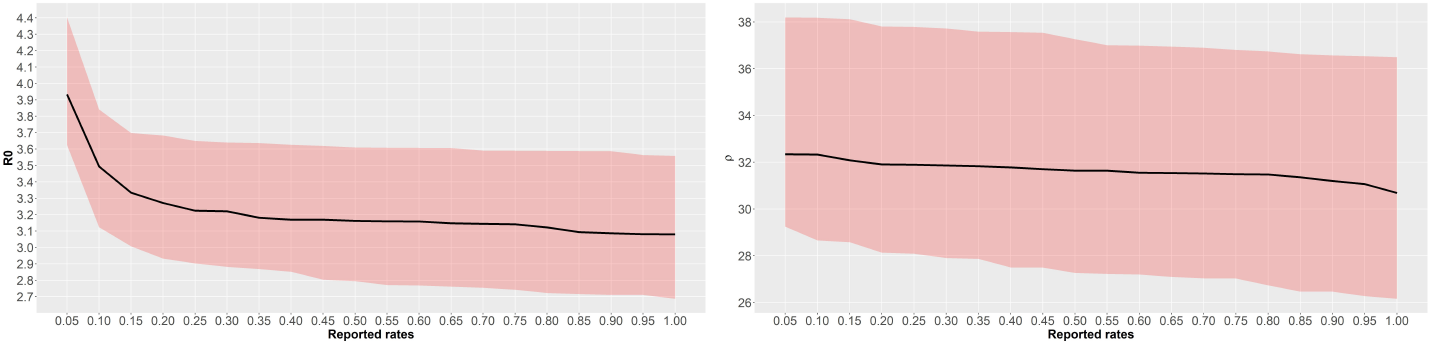
Point estimates and 95% credible intervals for *R*_0_ and infection period versus reported rates.

Figure 4 shows the estimated SIR curves for COVID-19 versus reported rate, from which we observe that the lower the reported rate, the earlier the peak is reached with a higher proportion of infected individuals. It is also observed that the contagion curves become similar to each other as the reported rates increase. These results reveal that the peak estimate of the COVID-19 transmission curve in Brazil is compromised when the presence of under-reporting is ignored.

**Figure 4:**
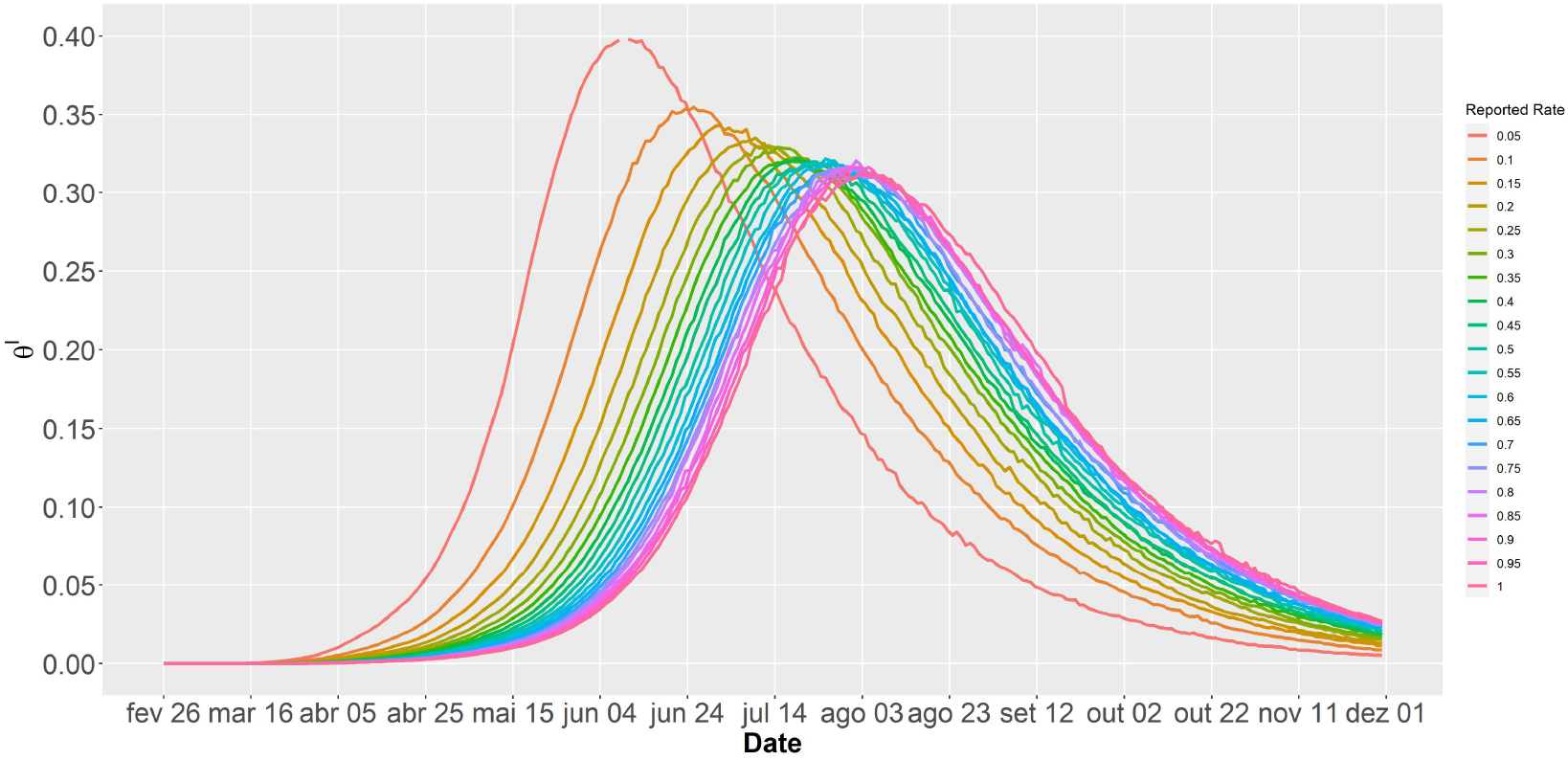
Estimated SIR curves versus reported rate for COVID-19 Brazilian data.

Finally, Table 4 presents the deviance information criterion (DIC) (Spiegelhalter et al., 2002), which indicates the SIR model with the reported rate of 0.1 as the best one that fitted the simulated data, since its DIC value is the lowest. These results suggest that the notification rate is very low.

**Table 4:** DIC values for COVID-19 Brazilian data.

| rate($\eta$) | DIC | rate($\eta$) | DIC |
| --- | --- | --- | --- |
| 0.05 | 3197.96 | 0.55 | 3260.65 |
| 0.10 | 3183.47 | 0.60 | 3266.44 |
| 0.15 | 3208.17 | 0.65 | 3272.19 |
| 0.20 | 3237.96 | 0.70 | 3277.76 |
| 0.25 | 3241.55 | 0.75 | 3279.27 |
| 0.30 | 3248.51 | 0.80 | 3283.36 |
| 0.35 | 3249.57 | 0.85 | 3295.20 |
| 0.40 | 3258.51 | 0.90 | 3296.90 |
| 0.45 | 3259.62 | 0.95 | 3306.84 |
| 0.50 | 3259.71 | 1.00 | 3307.88 |

## 7. Concluding Remarks

In this paper, we show that the method of adjusting cases by delay can be used to determine the reported rate of COVID-19 cases. Thus, it was possible that the rate of cases reported in Brazil is 0.07 and thus underestimates the real spreading of pandemic in the country.

Thus we proposed a SIR model with correction for under-reporting. The Bayesian approach is a feasible way to deal with the parameters inherent to the SIR model.

The methods reached convergence in the application with the Brazilian COVID-19 data set. Thus, a reproductive rate of 3.6243 was obtained, indicating that the epidemic is still booming in Brazil.

The simulation study revealed that the parameters estimates from the SIR model and the peak estimate which is a concern of several researchers and health authorities are sensitive to reporting rates. Future work may include considering the use of extended SIR models like the SEIR model (with the compartments of susceptible, exposed, infected, and removed individuals), and further, consider different scenarios of isolation and quarantine for the strategy of the COVID-19 transmission control.

## Data Availability

No Data Availability Statement

## Appendix A. Numerical Solution for SIR model

Let *f* (***θ****_t_*_−1_, *β*, *γ*) be the Runge-Kutta RK4 approximation to the SIR model.

Thus,

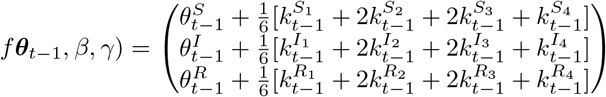

where

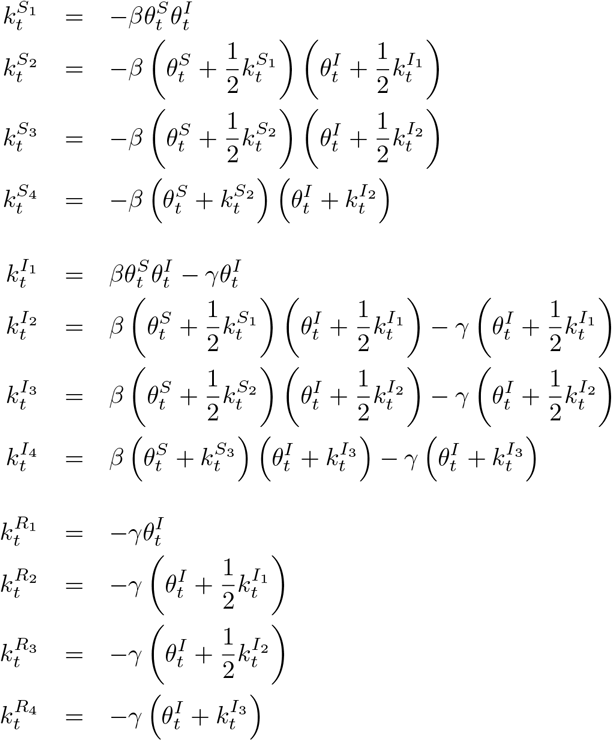

